# Explainable Advanced Electrocardiography Heart Age Shows Good Reproducibility in Healthy Young Adults

**DOI:** 10.64898/2026.03.24.26349147

**Authors:** Callum Warrington, Zaidon Al-Falahi, Upul Premawardhana, Martin Ugander, Simon Green

## Abstract

**Aims:** Explainable advanced electrocardiography (A-ECG) can be used to estimate heart age from the standard 12-lead ECG. A-ECG heart age gap (HAG) represents the difference between A-ECG heart age and chronological age. Increased A-ECG HAG is associated with cardiovascular outcomes and can be used to communicate risk. The aim was to investigate whether A-ECG heart age demonstrates acceptable within- and between-session reproducibility.

**Methods:** Healthy adults (n=42, age 23±4 years, 52% male) attended up to two sessions ~14 days apart, with 36 participants completing both sessions. During each session, five standard resting 12-lead ECGs were obtained while lying in the supine position with unchanged electrode positions. A-ECG heart age was extracted using dedicated software. Within-session reproducibility was assessed using all five recorded ECGs with coefficient of variation (CV) and a two-way random effects intraclass correlation coefficient (ICC). Between-session reproducibility was assessed using the first recorded ECG of each session with a paired t-test, CV and ICC. A further analysis assessed the reproducibility of the parameters used in the A-ECG heart age regression model.

**Results:** A-ECG heart age showed excellent within-session reproducibility in session one and two (both CV 5.8%, ICC 0.99). A-ECG heart age was slightly lower in session one than two (24.0±7.5 vs. 25.5±7.8 years, p=0.04) and showed good between-session reproducibility (CV 8.3%, ICC 0.84). All but one parameter used to estimate A-ECG heart age showed acceptable within- and between-session reproducibility (CV<10%).

**Conclusion:** A-ECG heart age demonstrates excellent within-session reproducibility and good between-session reproducibility in healthy young adults.

**LAY SUMMARY:** This study investigated whether advanced electrocardiography (A-ECG) heart age, an explainable heart age estimate derived from a standard 12-lead ECG, shows acceptable reproducibility when estimated repeatedly in healthy young adults.

- A-ECG heart age showed excellent within-session reproducibility across five ECGs recorded during each of two sessions approximately 14 days apart, with within-session coefficients of variation (CV) of 5.8%. When comparing the first recorded ECGs from each session, A-ECG heart age offered good between-session reproducibility demonstrating a CV of 8.3%.
- These findings show that A-ECG heart age is stable under standardised recording conditions in healthy young adults.

**Graphical abstract:** 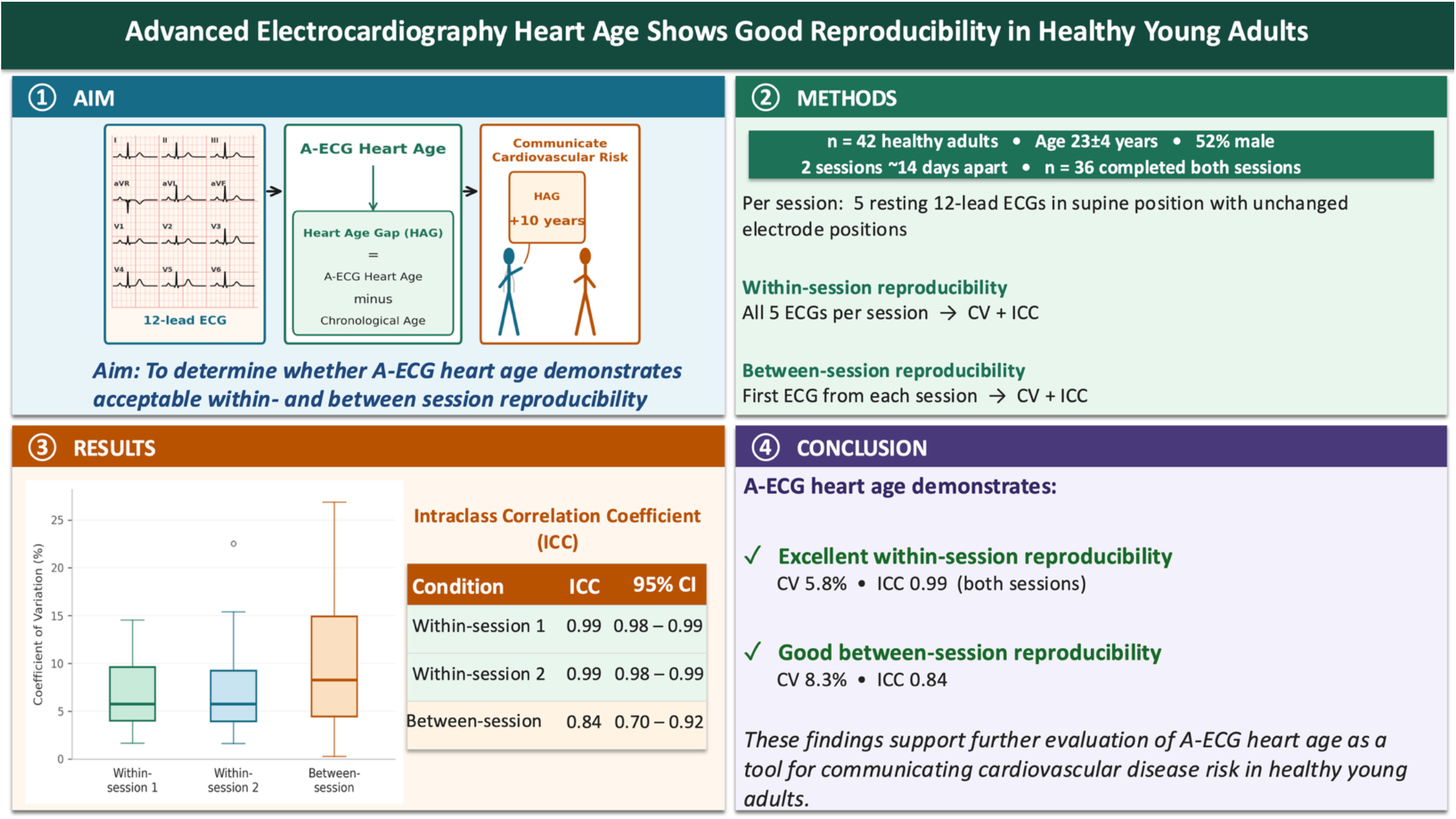

## Introduction

Recent evidence has shown that communication of a patient’s heart age can be a cost- and time-effective method for promoting positive lifestyle changes that reduce cardiovascular disease (CVD) risk ^1^. However, most heart age estimates are derived from online calculators, which have been shown to have a lack of agreement between models, often with inconsistent results ^2,3^.

Explainable advanced electrocardiography (A-ECG) is an emerging approach to analysing the standard 12-lead ECG, which generates ~500 parameters by combining conventional ECG features with those derived from the previously established techniques of vectorcardiography and waveform complexity analysis via singular value decomposition ^4^. Of these parameters, a subset is used in a multivariable regression model to estimate A-ECG heart age ^5^. Notably, the regression coefficients of the A-ECG parameters included in the heart age model differ between males and females to account for sex differences in the ECG ^5,6^. Importantly, A-ECG heart age originally required the presence of quantifiable P waves for its estimation ^7,8^. However, a recent refinement to the regression model now allows A-ECG heart age to be derived exclusively from QRS and T wave features, extending its applicability to both sinus and non-sinus rhythms ^5^.

A-ECG heart age gap (HAG) is the difference between A-ECG heart age and chronological age and is associated with increased risk of hospitalisation for heart failure and all-cause mortality ^5^. Across both sexes, healthy individuals demonstrate A-ECG HAGs of (mean±SD) 0.4±5.0 years, compared with 7.4±6.8 years in individuals with CVD risk factors and 13.5±8.3 years in patients with established CVD ^5^. Although large SDs would be expected in populations with CVD and risk factors due to condition heterogeneity, it remains unknown whether the variability among healthy individuals reflects CVD risk factors that were unaccounted for or measurement error.

To date, all studies of A-ECG heart age have relied on single standard 12-lead ECGs retrospectively obtained from hospital databases ^5,7,8^. As a result, the reproducibility of A-ECG heart age under prospectively standardised recording conditions has not yet been investigated ^5,7,8^. Before A-ECG heart age and HAG can be more broadly applied in CVD risk communication, acceptable within- and between-session reproducibility must be demonstrated ^5,9,10^.

We aim to use prospectively acquired, standardised 12-lead ECG recordings to assess the within- and between-session reproducibility of A-ECG heart age in healthy young adults. A secondary aim is to determine whether A-ECG HAG shows variance across sex, within-sessions, or between-sessions.

## Methods

### Participants

Forty-two healthy adult medical students (22 males and 20 females) participated in the study. Participants were screened for self-reported history of known CVD, cardiac symptoms and clinically abnormal ECG findings. If any participants were found to have clinically significant cardiac abnormalities, then they were referred to a cardiologist and excluded from the data analysis.

### Experimental Design

A repeated measures design was used with each participant attending two sessions separated by ~14 days. This interval was selected to reflect the time between follow-up visits in an outpatient cardiology clinic and to introduce variation in the menstrual cycle for female participants ^11,12^. During the first session, participants were asked to complete a standard cardiac health screening questionnaire. This was followed by assessment of height, weight and brachial artery blood pressure. For each session, five resting 12-lead ECG recordings were taken under standardised conditions. All recorded ECGs were uploaded to dedicated software for extraction of A-ECG heart ages. Ethics approval was obtained from South Western Sydney Local Health District Human Research Ethics Committee (ethics approval number: 2024/ETH00851) and all participants provided written informed consent.

### Procedures

For the first session only, participants completed a standard cardiac health screening questionnaire. This captured demographic data including sex, ethnicity, smoking status and family history of CVD. Additionally, body weight was measured using calibrated scales and recorded to the nearest 1 kg and height was measured to the nearest 1 cm using a stadiometer. Finally, resting left brachial artery pressure was measured using a digital sphygmomanometer (Omron HEM-7130, Omron Healthcare, Kyoto, Japan) while participants remained at rest in a seated position.

During each session, five resting 12-lead ECGs were recorded using a clinical ECG vendor (PageWriter TC50, Philips Healthcare, Australia) using standard electrode placement ^13^. Participants remained in a supine and rested position throughout the recordings. For repeated measurements within a session, electrodes were left unchanged for all ECG recordings. During the second session new electrodes were applied according to the same standard placement guidelines followed for the first session to maintain consistency ^13^.

### A-ECG and Statistical Analysis

All ECGs were analysed using dedicated A-ECG analysis software. The software extracted the parameters relevant to A-ECG heart age, which were then used in a multivariable regression model to generate A-ECG heart age estimates. This produced five A-ECG heart ages and corresponding HAGs for each session attended by participants.

Normality of data was checked using a Shapiro-Wilk test. Normally distributed data are presented as mean±SD and non-normally distributed data are presented as median and interquartile range (IQR). Descriptive statistics were used to summarise participant demographics and independent t tests were used to assess sex differences in baseline characteristics. All statistical analyses were conducted in R (version 4.2.2, R Foundation for Statistical Computing, Vienna, Austria), with significance level set at p<0.05.

For each session, within-session reproducibility for A-ECG heart age was assessed using the coefficient of variation (CV), a two-way random-effects, average-measures intraclass correlation coefficient (ICC) with 95% confidence intervals (CI) and minimum detectable change at 95% (MDC95). CV was calculated for each participant as (SD / mean × 100) across the five repeated A-ECG heart age estimates, with the group value presented as median [IQR]. If one or more ECG recordings obtained from a participant within a session could not be analysed by the A-ECG software, they were excluded from the ICC and MDC95 calculations, as these require a complete set of five measurements.

Between-session reproducibility for A-ECG heart age was assessed, using the first ECG recordings from each session to reflect a clinically realistic comparison between separate visits, with a paired t-test, CV and a two-way random-effects single-measure ICC with CI. Between-session CV was calculated for each participant as (SD / mean × 100) using the first ECG recording from each session, with the group value presented as median [IQR]. Agreement between-sessions for A-ECG heart age was further examined with Bland-Altman analysis to calculate the mean bias and 95% limits of agreement.

To complement these analyses, we also assessed the within- and between-session reproducibility of the individual parameters used to estimate A-ECG heart age with CV, calculated using the same per-participant approach described above. These A-ECG parameters have been described previously ^5^. For A-ECG parameters with negative sign, CV was calculated as 100×SD/|mean| to report a positive magnitude.

A three-way mixed repeated-measures analysis of variance (ANOVA) was performed to assess whether A-ECG HAG varied across the within-session HAGs, between-session HAGs and sex. For the ANOVA, only participants with all five ECG recordings successfully analysed in both sessions were included. Within-session HAGs and between-session HAGs were included as within-subject factors, while sex was included as a between-subject factor. A Greenhouse-Geisser correction was applied to effects involving within-session repeats.

## Results

### Participants

All 42 participants (22 males and 20 females) who attended session one had no cardiac abnormalities and were included in data analysis (Table 1). Of these, 36 participants (18 males and 18 females) attended both sessions, while the remaining six (4 males and 2 females) withdrew before session two. All 42 participants were included in the within-session reproducibility analysis for session one. The 36 participants who attended both sessions were included in the within-session reproducibility analysis for session two and the between-session reproducibility analysis. For the within-session ICC and MDC95 calculations, only participants with all five ECG recordings successfully analysed by the A-ECG software were included (session one: n=39, session two: n=34). For the ANOVA, only participants with all five recordings successfully analysed in both sessions were included (n=31, 14 males and 17 females).

**Table 1:** Participant Characteristics.

| Variable | Male (n=22) | Female (n=20) | p-value | Total (n=42) |
| --- | --- | --- | --- | --- |
| Chronological age, years | 22.4 ± 3.3 | 23.5 ± 4.0 | 0.34 | 22.9 ± 3.6 |
| BMI, kg/m<sup>2</sup> | 25.0 ± 2.5 | 21.9 ± 3.2 | **0.001** | 23.5 ± 3.2 |
| Height, cm | 178 ± 9 | 165 ± 6 | **<0.001** | 171 ± 10 |
| Weight, kg | 79 ± 13 | 59 ± 9 | **<0.001** | 70 ± 15 |
| SBP, mmHg | 131 ± 14 | 112 ± 10 | **<0.001** | 122 ± 16 |
| DBP, mmHg | 79 ± 9 | 76 ± 9 | 0.21 | 78 ± 9 |
| MAP, mmHg | 96 ± 8 | 88 ± 9 | **0.002** | 92 ± 9 |

Data are presented as mean ± SD. BMI = body mass index, SBP = systolic blood pressure, DBP = diastolic blood pressure, MAP = mean arterial pressure.

### Within-Session Reproducibility

A-ECG heart age showed excellent within-session reproducibility in both session one (CV 5.8% [4.0%, 9.6%], ICC 0.99 (95%CI 0.98 to 0.99), MDC95 5.0 years) and two (CV 5.8% [3.9%, 9.2%] and ICC 0.99 (95%CI 0.98 to 0.99), MDC95 5.4 years). Consistent with these findings, all but one of the A-ECG parameters used for heart age estimation demonstrated within-session CV<10% (Figure 1).

**Figure 1.**
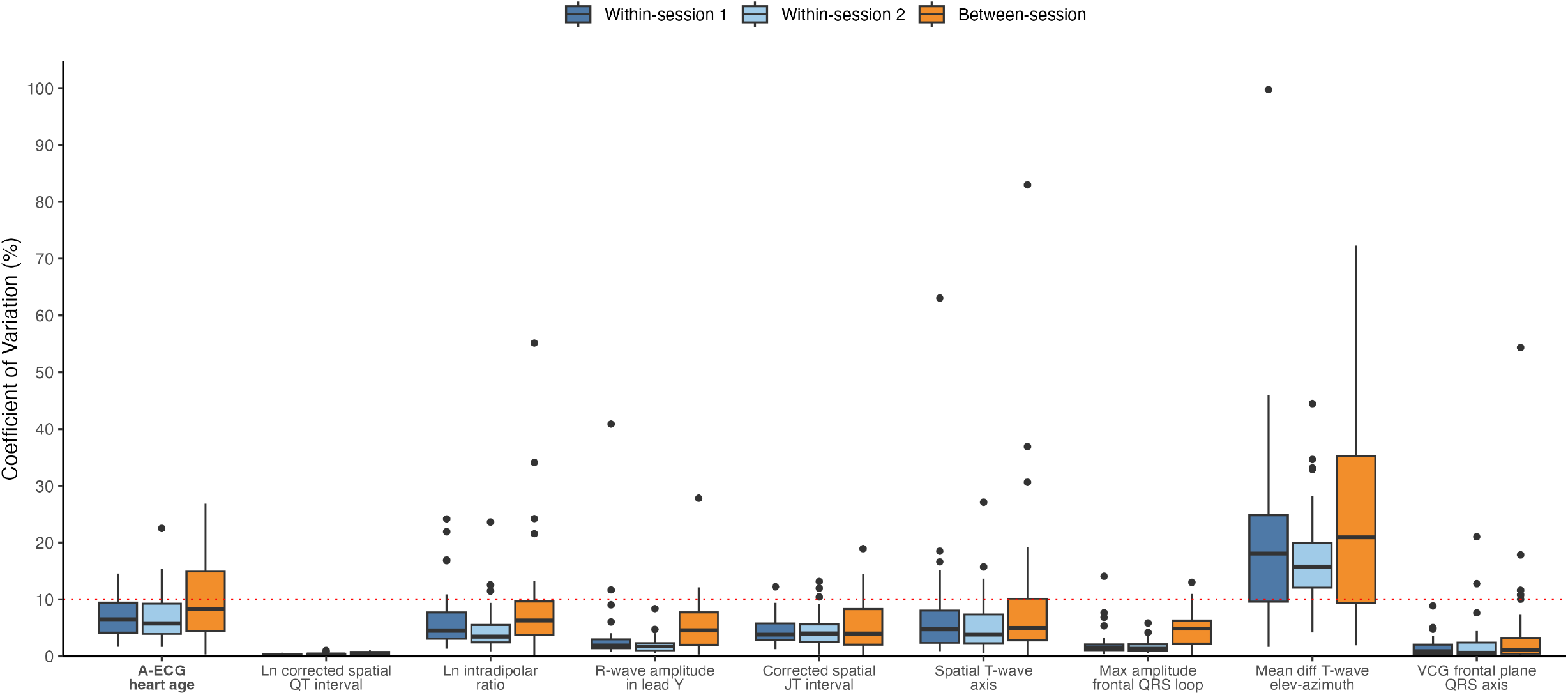
Within- and between-session coefficient of variations (%) for A-ECG heart age and the individual parameters used in the A-ECG heart age multivariable regression model. Data are displayed as boxplots, where the horizontal line represents the median, the box represents the interquartile range, whiskers extend to the most extreme values within 1.5 times the interquartile range, and points beyond the whiskers represent individual outliers. The red dotted line indicates a CV of 10%. For the ln-transformed intradipolar ratio of T-wave complexity from the Frank XYZ leads [(T Eigenvalue2 × T Eigenvalue3)/(T Eigenvalue1)^2^] following singular value decomposition, CV was calculated as 100 × SD/|mean| to ensure a positive magnitude, as ln-transformed values are negative.

**Figure 2.**
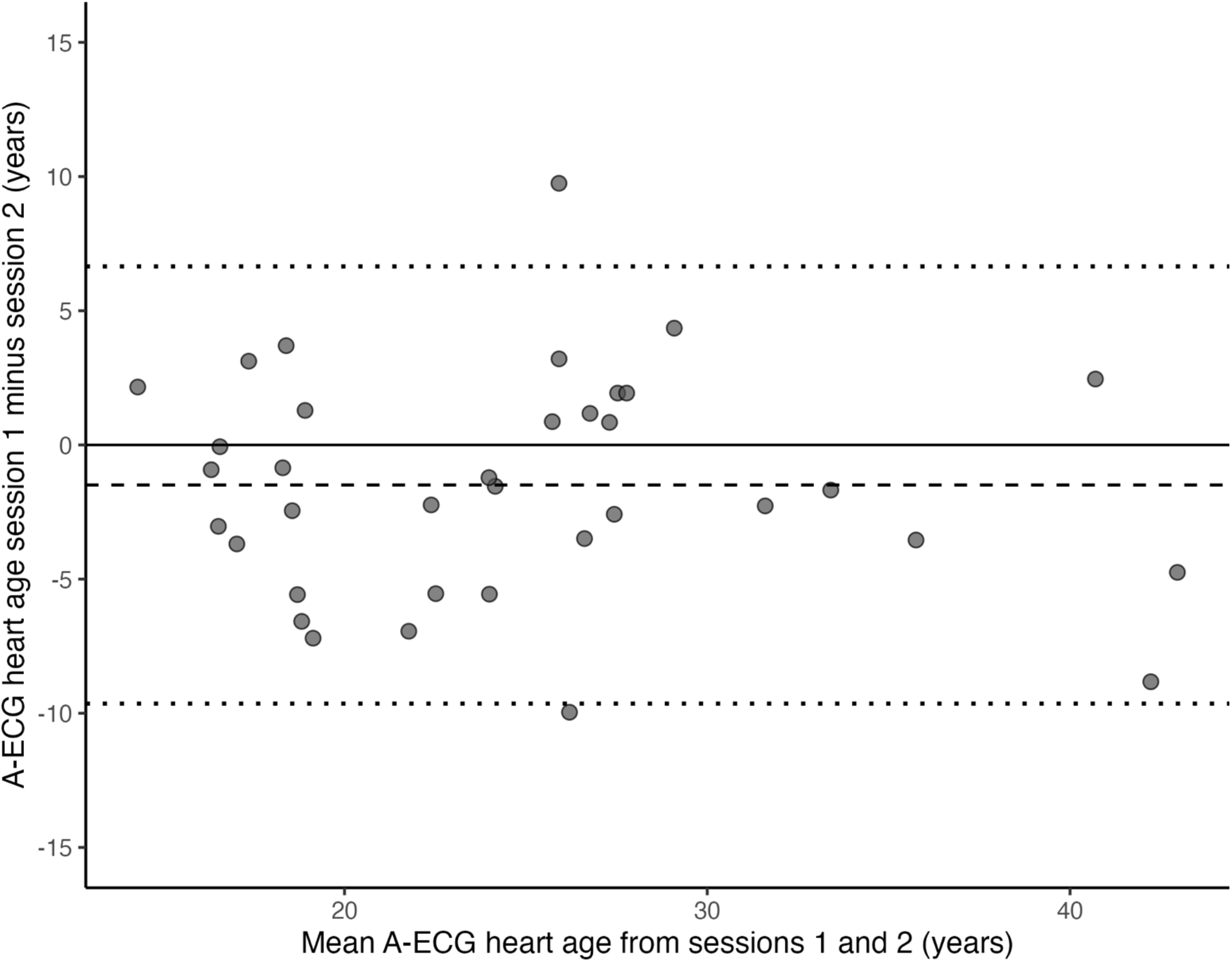
Bland-Altman plot of between-session agreement for A-ECG heart age. Each data point represents one participant (n = 36). The y-axis is the difference in A-ECG heart age between session 1 and session 2 (years), the x-axis is the mean of the two sessions (years). The dashed line indicates the mean bias (−1.5 years). Dotted lines indicate the 95% limits of agreement (upper: +6.7 years; lower: −9.6 years), calculated as bias ± 1.96 x SD of the pairwise differences.

### Between-Session Reproducibility

The first acquired A-ECG heart ages in each session were lower in session one than session two (24.0±7.5 vs. 25.5±7.8 years, p=0.04). However, good between-session reproducibility was observed (CV 8.3% [4.5%, 14.9%], ICC 0.84 (95%CI 0.70 to 0.92)). Bland-Altman analysis revealed a mean bias of −1.5±4.2 years and the limits of agreement ranged from −9.6 to +6.7 years. Consistent with these findings, all but one of the A-ECG parameters used for heart age estimation demonstrated between-session CV<10% (Figure 1).

### Effects of within-session HAGs, between-session HAGs and sex

Across the three main factors of within-session HAGs, between-session HAGs and sex, there was no significant effect of within-session A-ECG HAGs (p=0.79). However, there was a significant effect of between-session HAGs, with lower A-ECG HAGs observed in session one than session two (1.1±6.1 vs. 2.2±6.5 years, p=0.03). There was no effect of sex (p=0.70), with males and females demonstrating similar A-ECG HAGs (1.5±4.8 vs. 1.8±7.5 years). A significant interaction between session and within-session repeats was also observed (p=0.03), though the effect size was small (generalised η^2^=0.003). No other interactions were significant (p>0.05 for all).

## Discussion

In this first investigation of A-ECG heart age using prospectively acquired and fully standardised ECG recordings, A-ECG heart age showed ICC values signifying excellent within-session reproducibility and good between-session reproducibility ^9^. Within-session reproducibility was further supported by low CVs and small MDC95 values relative to known differences in A-ECG HAG between healthy individuals and those with CVD risk factors ^5^. Between-session reproducibility was similarly supported by only a marginally higher CV compared with within-session values, and Bland-Altman analysis revealed a small mean bias between sessions. Although A-ECG heart age was significantly lower in session one than session two, this difference of 1.5 years was small in magnitude. Consistent with these findings, CV values below 10% were observed both within and between sessions for all but one of the parameters used to estimate A-ECG heart age. Finally, A-ECG HAG showed variance between-session, but not within-session or across sex.

The within-session ICC for A-ECG heart age in both sessions was indicative of excellent reproducibility ^9^. Furthermore, the low CV values indicate that the variability of A-ECG heart age within sessions was small and is comparable to what has been reported in other measures of CVD risk such as automated sphygmomanometry ^14^ and continuous glucose monitoring ^15^. Importantly, the MDC95 values suggest that only changes in A-ECG heart age smaller than 5.0 to 5.4 years may be attributable to measurement error. These MDC95 values are smaller than the differences previously observed between healthy individuals and those with CVD risk factors or established CVD, where mean A-ECG HAGs rise from approximately 0 years to 7 to 14 years ^5,8^.

The between-session ICC for A-ECG heart age was indicative of good reproducibility ^9^. Visual inspection of the Bland-Altman plot also revealed a small mean bias between sessions and the between-session CV of 8.3% was only marginally higher than the within-session CVs of 5.8%. Notably, the upper limit of agreement of +6.7 years closely aligns with the upper range of A-ECG HAGs found in healthy individuals ^5^.

In support of these findings, the individual parameters used to estimate A-ECG heart age showed acceptable reproducibility (CV<10%) both within- and between-sessions, with the exception of the mean difference between the frontal plane T-wave elevation and azimuth of all T-wave loop samples, which exceeded a CV of 10%. However, due to this parameter having the smallest standardised t ratio in the multivariable regression model, it did not compromise the overall reproducibility of the A-ECG heart age estimate ^5^. This is consistent with prior conventional ECG reproducibility research which showed that, although some individual ECG measurements can exhibit relatively large CVs compared to others, this does not necessarily translate into clinical reclassifications when interpreting multiple ECG recordings ^16^.

The ANOVA findings further support the reproducibility of A-ECG heart age, as A-ECG HAGs did not show significant variance within each session. This suggests that a single ECG recording may provide a reliable estimate of A-ECG HAG in healthy adults under highly standardised conditions. Although there was a statistically significant difference in A-ECG HAG between sessions, the difference was only 1.1 years, which is a small magnitude. While the interaction between within-session HAGs and between-session HAGs was significant, the effect size was negligible.

The absence of a significant effect of sex on A-ECG HAG suggests that it is similar in males and females. This provides support for the sex-specific regression models used to estimate A-ECG heart age, indicating that they appropriately account for physiological sex differences in cardiac electrical activity in healthy young adults ^5,8^. This contrasts with other markers of CVD risk, such as coronary artery calcium scores and left ventricular mass index, where clinical interpretation can vary according to sex ^17,18^.

The current findings have several potential clinical implications. First, the excellent within-session reproducibility suggests that a single ECG recording may be sufficient to provide a reliable estimate of A-ECG heart age under highly standardised conditions. This supports further evaluation of A-ECG heart age within outpatient cardiology clinics, where a single ECG recording is usually obtained at each visit ^11^. Second, the good between-session reproducibility supports further investigation of A-ECG heart age as a measure that may be suitable for tracking changes in CVD risk over time. This may be relevant in future research examining the impact of lifestyle modification or pharmacological intervention on A-ECG HAG ^5,19,20^. Third, the relatively small measurement error observed in this study suggests that substantial increases in A-ECG HAG over time may be more likely to reflect clinically meaningful changes in CVD risk. This supports further evaluation of A-ECG heart age as a potential marker of deviation from healthy cardiac ageing ^5,21^.

Several limitations should be considered when interpreting these findings. First, the sample consisted of healthy young adults, which limits generalisability to older populations and patients with CVD. Second, all ECGs were recorded by a single researcher using standardised electrode placement. Therefore, reproducibility may be lower in hospital and outpatient settings where ECGs are acquired by multiple clinical staff and electrode placement may vary. Finally, all ECGs were recorded using the same vendor, thus these findings may not directly translate across different manufacturers.

## CONCLUSION

In summary, A-ECG heart age showed excellent within-session reproducibility and good between-session reproducibility under standardised conditions in healthy young adults. Additionally, A-ECG HAG did not differ by sex. These findings support further evaluation of A-ECG heart age as a tool for communicating CVD risk in healthy young adults of both sexes ^5^.

## Data Availability

All data produced in the present study are available upon reasonable request to the authors

## ACKNOWLEDGMENTS

The authors thank Western Sydney University medical students, Andreas Feiden, Antonius Yasintus, Claudia Tiong and David Jiawei Ou, for their assistance in participant recruitment in this study.

## DATA AVAILABILITY

The datasets generated and analysed during the current study are available from the corresponding author on reasonable request.

## REFERENCES

1. Lopez-Gonzalez AA, Aguilo A, Frontera M, Bennasar-Veny M, Campos I, Vicente-Herrero T, et al. ECectiveness of the Heart Age tool for improving modifiable cardiovascular risk factors in a Southern European population: a randomized trial. Eur J Prev Cardiol 2015;22:389–396. doi: 10.1177/2047487313518479

2. Bonner C, Bell K, Jansen J, Glasziou P, Irwig L, Doust J, et al. Should heart age calculators be used alongside absolute cardiovascular disease risk assessment? BMC Cardiovascular Disorders 2018;18:19. doi: 10.1186/s12872-018-0760-1

3. Bonner C, Fajardo MA, Hui S, Stubbs R, Trevena L. Clinical Validity, Understandability, and Actionability of Online Cardiovascular Disease Risk Calculators: Systematic Review. J Med Internet Res 2018;20:e29. doi: 10.2196/jmir.8538

4. Schlegel TT, Kulecz WB, Feiveson AH, Greco EC, DePalma JL, Starc V, et al. Accuracy of advanced versus strictly conventional 12-lead ECG for detection and screening of coronary artery disease, left ventricular hypertrophy and left ventricular systolic dysfunction. BMC Cardiovasc Disord 2010;10:28. doi: 10.1186/1471-2261-10-28

5. Al-Falahi ZS, Schlegel TT, Palencia-Lamela I, Li A, Schelbert EB, Niklasson L, et al. Advanced electrocardiography heart age: a prognostic, explainable machine learning approach applicable to sinus and non-sinus rhythms. European Heart Journal - Digital Health 2024. doi: 10.1093/ehjdh/ztae075

6. St Pierre SR, Peirlinck M, Kuhl E. Sex Matters: A Comprehensive Comparison of Female and Male Hearts. Front Physiol 2022;13:831179. doi: 10.3389/fphys.2022.831179

7. Lindow T, Maanja M, Schelbert EB, Ribeiro AH, Ribeiro ALP, Schlegel TT, et al. Heart age gap estimated by explainable advanced electrocardiography is associated with cardiovascular risk factors and survival. Eur Heart J Digit Health 2023;4:384–392. doi: 10.1093/ehjdh/ztad045

8. Lindow T, Palencia-Lamela I, Schlegel TT, Ugander M. Heart age estimated using explainable advanced electrocardiography. Scientific Reports 2022;12:9840. doi: 10.1038/s41598-022-13912-9

9. Atkinson G, Nevill AM. Statistical methods for assessing measurement error (reliability) in variables relevant to sports medicine. Sports Med 1998;26:217–238. doi: 10.2165/00007256-199826040-00002

10. Batterham AM, George KP. Reliability in evidence-based clinical practice: a primer for allied health professionals. Physical Therapy in Sport 2003;4:122–128. doi: 10.1016/S1466-853X(03)00076-2

11. Bots SH, Siegersma KR, Onland-Moret NC, Asselbergs FW, Somsen GA, Tulevski, II, et al. Routine clinical care data from thirteen cardiac outpatient clinics: design of the Cardiology Centers of the Netherlands (CCN) database. BMC Cardiovasc Disord 2021;21:287. doi: 10.1186/s12872-021-02020-7

12. McKinley PS, King AR, Shapiro PA, Slavov I, Fang Y, Chen IS, et al. The impact of menstrual cycle phase on cardiac autonomic regulation. Psychophysiology 2009;46:904–911. doi: 10.1111/j.1469-8986.2009.00811.x

13. Campbell B, Richley D, Ross C, Eggett CJ. Clinical Guidelines by Consensus: Recording a standard 12-lead ECG. https://scst.org.uk/wp-content/uploads/2024/09/2024_ECG_Recording_Guidelines_26-09-2024_V5_FINAL.pdf (31 May 2025)

14. Stanforth PR, Gagnon J, Rice T, Bouchard C, Leon AS, Rao DC, et al. Reproducibility of Resting Blood Pressure and Heart Rate Measurements: The HERITAGE Family Study. Annals of Epidemiology 2000;10:271–277. doi: 10.1016/S1047-2797(00)00047-8

15. Matabuena M, Pazos-Couselo M, Alonso-Sampedro M, Fernández-Merino C, González-Quintela A, Gude F. Reproducibility of continuous glucose monitoring results under real-life conditions in an adult population: a functional data analysis. Scientific Reports 2023;13:13987. doi: 10.1038/s41598-023-40949-1

16. McLaughlin SC, Aitchison TC, Macfarlane PW. The value of the coeCicient of variation in assessing repeat variation in ECG measurements. Eur Heart J 1998;19:342–351. doi: 10.1053/euhj.1997.0741

17. Bigeh A, Shekar C, Gulati M. Sex DiCerences in Coronary Artery Calcium and Long-term CV Mortality. Curr Cardiol Rep 2020;22:21. doi: 10.1007/s11886-020-1267-9

18. Salton CJ, Chuang ML, O’Donnell CJ, Kupka MJ, Larson MG, Kissinger KV, et al. Gender diCerences and normal left ventricular anatomy in an adult population free of hypertension: A cardiovascular magnetic resonance study of the Framingham Heart Study OCspring cohort. Journal of the American College of Cardiology 2002;39:1055–1060. doi: 10.1016/S0735-1097(02)01712-6

19. Kariuki JK, Imes CC, Engberg SJ, Scott PW, Klem ML, Cortes YI. Impact of lifestyle-based interventions on absolute cardiovascular disease risk: a systematic review and meta-analysis. JBI Evid Synth 2024;22:4–65. doi: 10.11124/jbies-22-00356

20. Cicero AFG, Fogacci F, Rizzoli E, Giovannini M, D’Addato S, Borghi C. Impact of combined treatment with ACE inhibitors and statins on cardiovascular outcomes in the Brisighella Heart Study: A 8-year Follow-Up. Journal of Hypertension 2022;40:e213. doi: 10.1097/01.hjh.0000837632.68874.6e

21. Vriend EMC, Artola Arita V, Menassa M, Beigrezaei S, Moll van Charante EP, van den Born B-JH, et al. Cardiovascular ageing definition: a scoping review on conceptual and operational frameworks. European Journal of Preventive Cardiology 2025. doi: 10.1093/eurjpc/zwaf741

